# Domain-specific Cognitive Impairments, Mood and Quality of Life 6 Months After Stroke

**DOI:** 10.1101/2024.01.24.24301716

**Authors:** Elise Milosevich, Andrea Kusec, Sarah T. Pendlebury, Nele Demeyere

## Abstract

**Purpose:** To identify which acute and 6-month domain-specific cognitive impairments impact mood functioning, participation, and stroke-related quality of life 6 months after stroke.

**Materials and Methods:** A prospective cohort of 430 stroke survivors completed the Oxford Cognitive Screen (OCS) acutely and 6 months post-stroke. Participants completed the Stroke Impact Scale (SIS) and Hospital Depression and Anxiety Scale (HADS) at 6 months. Multivariable regression analyses assessed whether severity of, and domain-specific, cognitive impairment acutely and at 6 months was associated with composite 6-month SIS scores, each SIS subscale, and HADS scores.

**Results:** Increased severity of cognitive impairment acutely and at 6 months was associated with lower 6-month SIS composite scores independent of age, sex, education years, and stroke severity (both *p*<0.001). Domain-specific impairments in memory (*p*<0.001) and attention (*p*=0.002) acutely, and language (*p*<0.001), memory (*p*=0.001) and number processing (*p*=0.006) at 6 months showed the strongest associations with worse SIS composite scores. Severity of acute and 6-month cognitive impairment was associated with poorer functioning in each SIS subscale, as well as greater levels of depression (acute *p*=0.021, 6-months *p*<0.001), but not anxiety (*p*=0.174, *p*=0.129).

**Conclusions:** Both acute and 6-month domain-specific cognitive impairments, particularly in memory, were found to negatively impact overall functional and mood outcomes 6 months post-stroke.

## INTRODUCTION

Stroke survivors commonly experience cognitive^1,2^ and emotional problems^3,4^, which can greatly impact reintegration into pre-stroke social and occupational roles, often limiting meaningful participation in activities of daily living (ADLs).^5,6^ Community-dwelling stroke survivors report numerous long-term unmet care needs,^7–9^ most frequently in the areas of cognition and emotion,^7^ followed by ADLs and participation.^9^ Consequently, the lack of services provided and care pathways available leave stroke survivors often feeling abandoned by health organizations and professionals when returning to the community.^8^ Longer-term monitoring of cognitive and emotional functioning could identify psychological factors which most influence overall quality of life after stroke. This could lead to improved facilitation of community reintegration.

Cognitive impairment has been associated with post-stroke depression, activities of daily living, and participation.^4,6,10^ However, a recent meta-analysis highlighted that the majority of studies investigating the relationship between cognition and functional outcomes have used a brief global cognitive screen, with very few studies reporting data on more than one cognitive domain.^6^ A stroke can result in impairment in different cognitive domains, therefore it is critical to consider all domains when determining how cognition impacts other post-stroke outcomes.^4,11–13^ There is currently an absence of consistent prevalence estimates of domain-specific cognitive impairments following stroke, resulting in a lack of understanding of how persistent domain-specific cognitive impairments influence the quality of life of stroke survivors.

Emotional functioning is another crucial determinant of post-stroke quality of life.^14^ The prevalence of mood disorders after stroke is considerably higher than in the general population,^15^ with depression being most prevalent ranging between 18-33%,^16–18^ while anxiety rates range from 19%-24%.^19^ Though both depression and anxiety have been shown to reduce participation,^20,21^ it is less clear whether cognitive impairment is linked to mood functioning following stroke. Global cognitive function has been shown to be an important predictor of post-stroke depression,^22^ however reported domain-specific cognitive correlates have been inconsistent.^4,16,23^ In comparison to post-stroke depression, the association between post-stroke anxiety and cognition (both global and domain-specific) has received far less attention.^16,24^ Furthermore, despite their frequent co-existence, depression and anxiety are rarely considered together following stroke.^16,24^

As stroke can result in multiple functional limitations, measures that capture a variety of recovery outcomes should be prioritized.^25^ Very few studies have investigated the association between cognition and multiple functional outcomes^13,26^ (e.g., ADLs, instrumental ADLs and participation) and instead have often focused on physical disability (e.g., with the Barthel Index or Modified Rankin Scale).^27^ Though post-stroke cognitive impairment has been shown to negatively impact both ADLs and participation,^6^ no studies to date have examined the contribution of domain-specific cognitive impairment to quality of life as measured by a stroke-specific multi-outcome measure; namely, the Stroke Impact Scale (SIS).^28^

This study sought to utilize a domain-specific cognitive screen, a stroke-specific multi-outcome measure, as well as a mood functioning measure which considers both depression and anxiety, in order to evaluate cognitive determinants of post-stroke quality of life. The primary aim was to assess the relationship between acute, as well as 6-month, severity of cognitive impairment and quality of life 6 months post-stroke measured by the SIS. This was further explored by examining the relationship between severity of acute and 6-month cognitive impairment and five SIS components (memory and thinking, communication and language, emotional functioning, activities of daily living, and participation), as well as depression and anxiety as measured by the Hospital Anxiety and Depression Scale (HADS).^29^ The secondary aim was to determine the relationship between domain-specific cognitive impairments and overall quality of life.

## MATERIALS AND METHODS

### Participants

A consecutive sample of stroke patients was recruited through the Oxford Cognitive Screening programme based within the John Radcliffe Hospital, UK acute stroke unit from 2012-2019. The present study considered existing data from a cohort of acute stroke survivors recruited between 2012-2019 (National Research Ethics Committee (UK) approval references 14/LO/0648 and 18/SC/0550). Participants were included if they had confirmed stroke by neuroimaging or clinical judgement, were aged 18 years or older and had sufficient English language comprehension to understand assessment instructions. Participant were excluded if they were unable to give written or witness informed consent or were unable to concentrate for 20 minutes as judged by the multidisciplinary team. A total of 866 stroke patients were recruited and received domain-specific cognitive screening acutely (within 2 weeks of admission), 430 of which completed follow-up assessment at 6 months (attrition details outlined in Supplementary figure 1). Detailed patient demographics, comorbidities, vascular risk factors and stroke characteristics were determined on admission through medical records and neuroimaging. Stroke severity was evaluated using the National Institutes of Health Stroke Scale^30^ (NIHSS).

### Cognitive Assessment

Domain-specific cognitive screening was carried out acutely (≤2 weeks) and at 6-months post-stroke using the Oxford Cognitive Screen (OCS).^31^ In this study, we used 12 metrics derived from the OCS subtests that are categorized into 6 cognitive domains: language (picture naming, semantic understanding, sentence reading), spatial attention (egocentric attention, allocentric attention), executive function (trail-making), memory (orientation, verbal memory, episodic memory), praxis (gesture imitation), and number processing (calculation and number writing).

Tasks were binarized into impaired or unimpaired based on published normative scores for each subtest.^31^ A domain impairment was characterized as at least one impaired task in that domain. Severity of impairment was characterized by the proportion of tasks impaired across all OCS domains. The OCS was administered by trained neuropsychologists or occupational therapists at bedside acutely and in-person follow-up. See Supplemental Methods for further considerations regarding subtests when administering the OCS.

### Functional and Quality of Life Assessment

The Stroke Impact Scale (SIS)*-Version 3*^28^ is a stroke-specific, self-report questionnaire and was used to assess overall functional ability and quality of life at 6-month follow-up only. The SIS comprises 59 items split across 8 subscales. The present study only focused on 5 of the SIS subcales: Memory and Thinking, Mood and Emotion, Communication and Language, ADLs, and Participation. The three subscales relating to the physical impact of stroke (i.e., Physical Strength, Mobility, and Hand Function) were not considered as they were not pertinent to the main research question and to reduce the number of multiple comparisons. Items are rated on a 5-point Likert scale, with higher scores indicating better function. Raw subscale scores are transformed into scores ranging from 0–100 using Equation 1.^32^ A single overall score from the 5 SIS domains was created through SIS Index methodology.^33^ This overall score was subsequently scaled using the equation below.

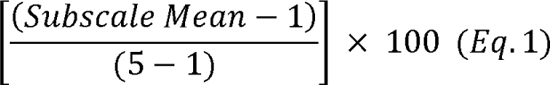

### Anxiety and Depression Assessment

Depression and anxiety symptomatology were assessed using the Hospital Anxiety and Depression Scale^29^ (HADS) at 6 months post-stroke only. The HADS consists of 7 items each for depression and anxiety with answers rated 0–3 for each question. Each subscale has a range of 0–21 for depression (HADS-D) and anxiety (HADS-A). Scores greater than 7 indicate possible cases of depression or anxiety, while scores greater than 10 indicate probable cases. Cut-off scores were used to describe the prevalence of anxious and depressive symptoms, whereas continuous HADS subscale scores were used for analyses.

### Statistical Analyses

Descriptive statistics were used to summarize sample demographics and clinical characteristics. Following this, a series of multivariable linear regressions were performed. The primary models examined the relationship between severity of cognitive impairment (proportion of tasks impaired) acutely and 6 months post-stroke and composite SIS scaled scores described above,^33^ as well as self-reported depression and anxiety severity on the HADS. Secondary models assessed the relationship between severity of acute and 6-month cognitive impairment with each individual SIS subscale score (i.e., instead of the composite score). The final model examined the relationship between impairment in each of the six individual cognitive domains acutely and at 6-months with overall quality of life, a single composite SIS scaled score. All regressions covaried for age, sex, years of education, and stroke severity (acute NIHSS scores). For all models, the effect of removing cases with high levels of influence as defined by a Cook’s distance^34^ was examined, however no significant difference was found, hence the full sample was used. An alpha of 0.05 was used as the cut-off for determining statistical significance. Dunn-Sidak corrections (alpha-adjusted 0.01) were employed where multiple analyses on the same dependent variable were conducted and reported where appropriate. All analyses were carried out in *R* version 4.2.0. The data which support the findings are available from the corresponding author upon reasonable request.

## RESULTS

A total of 430 stroke survivors (mean age 73.9 (SD 12.5) years, 46.5% female, median NIHSS 5; range 0–30) were assessed acutely after stroke (mean/SD 4.39/4.46 days) and again at mean/SD 6.65/1.06 months (table 1). A between-group comparison of demographics and acute cognitive function of those who did and did not complete 6-month follow-up, as well as the prevalence of domain-specific impairments, mean SIS subscale scores, and the number of cases of depression and anxiety is outlined in the Supplementary Materials (S1).

**Table 1.** Cohort demographics and acute clinical characteristics.

|  | **N = 430** |
| --- | --- |
| Age at stroke, mean (SD) | 73.86 (12.51) |
| Sex (female), n (%) | 200 (46.51) |
| Education years, mean (SD) | 12.25 (3.55) |
| Handedness, right n (%) | 374 (86.98) |
| *Stroke subtype, n (%)* |  |
| Ischaemic | 262 (84.19) |
| Haemorrhagic | 65 (15.12) |
| Mixed | 3 (1.00) |
| *Lesion side, n (%)* |  |
| Left | 153 (35.58) |
| Right | 168 (39.07) |
| Bilateral | 34 (7.91) |
| Undetermined* | 75 (17.44) |
| First-ever stroke, n (%) | 292 (67.91) |
| NIHSS, median [IQR]<sup>†</sup> | 5 [2–10] |
| NIHSS ≤ 3, n (%) | 121 (37.81) |
| NIHSS >3, n (%) | 199 (62.19) |
| Modified Rankin Scale, median [IQR]<sup>‡</sup> | 1 [0–2] |
| Independent at admission, n (%) | 376 (87.44) |
| Dependent (care required), n (%)<sup>§</sup> | 54 (12.56) |
| *Comorbidities, n (%)* |  |
| Atrial fibrillation | 109 (25.35) |
| Hypertension | 261 (60.70) |
| Diabetes mellitus | 83 (19.30) |
| *Smoking, n (%)* |  |
| Current | 44 (10.23) |
| Past | 34 (7.91) |
| Never | 352 (81.86) |
| Length of stay in hospital, mean days (SD) | 11.64 (11.44) |
| *Undetermined on admission acute CT scan |  |
| <sup>†</sup>NIHSS: National Institute of Health Stroke Scale (≤3 minor, >3 major) was recorded for n=320 (74%) |  |
| <sup>‡</sup>Modified Rankin Scale (mRS) (premorbid) was reported for n=248 (58%) |  |
| <sup>§</sup>Dependence was categorized as requiring formal carer support (by family n=18, by carer n=36) |  |

### SIS composite and subscales outcome

The distribution of the scaled self-reported SIS scores is visualised in figure 1. The mean composite SIS score was 72.77 (SD 20.48). Participants self-reported the highest functioning in communication, followed by memory, ADLs, emotions, and participation.

**Figure 1.**
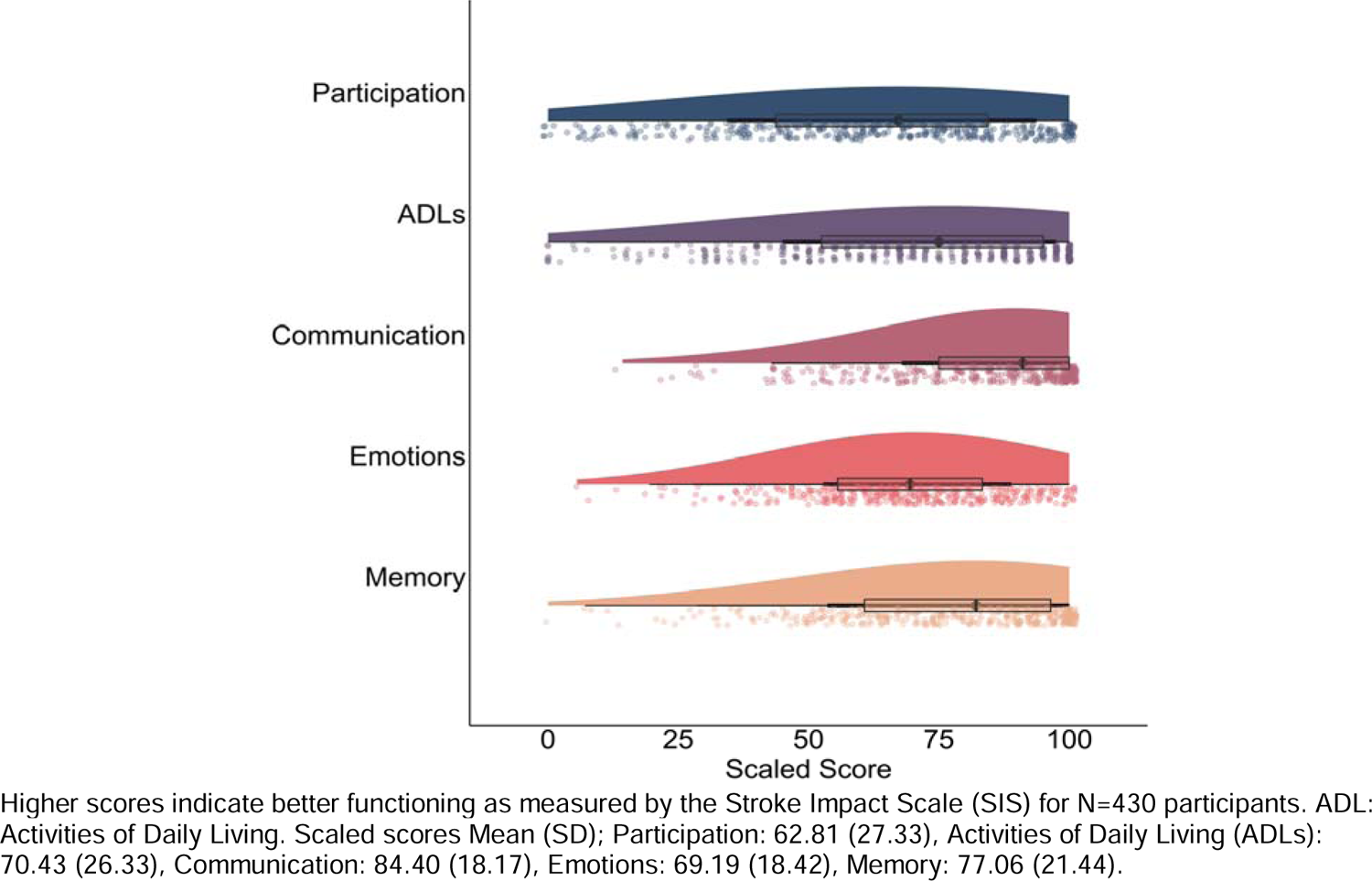
Distribution of the Stroke Impact Scale subscale scores (scaled) at 6 months post-stroke.

### Acute and 6-month cognitive impairment and quality of life at 6 months

Acutely (≤2 weeks) after stroke, increased severity of cognitive impairment was associated with lower 6-month SIS composite scores after adjusting for age, sex, years of education, and stroke severity (Adj R^2^=0.10, *p*=0.001). Increased severity of acute cognitive impairment was also associated with poorer self-reported functioning at 6 months in each of the SIS subscales after adjusting for covariates, except for the participation subscale (Adj R^2^=0.02, *p*=0.247). An overview of the regression results is outlined in table 2.

**Table 2.** Association between severity of acute and 6-month cognitive impairment and 5 components of quality of life.

| Dependent | SIS<br>Memory &<br>Thinking |  |  | SIS<br>Mood &<br>Emotions |  |  | SIS<br>Communication &<br>Language |  |  | SIS<br>Activities of<br>Daily Living |  |  | SIS<br>Participation |  |  |
| --- | --- | --- | --- | --- | --- | --- | --- | --- | --- | --- | --- | --- | --- | --- | --- |
| | b (SE) | $\beta$ (SE) | *t* | b (SE) | $\beta$ (SE) | *t* | b (SE) | $\beta$ (SE) | *t* | b (SE) | $\beta$ (SE) | *t* | b (SE) | $\beta$ (SE) | *t* |
| **Predictor** |  |  |  |  |  |  |  |  |  |  |  |  |  |  |  |
| Acute cognitive impairment | -27.34 (4.66) | -0.34 (0.06) | **-5.87\*\*\*** | -9.73 (4.23) | -0.14 (0.06) | **-2.30\*†** | -25.77 (3.74) | -0.39 (0.06) | **-6.89\*\*\*** | -12.70 (5.68) | -0.13 | **-2.24\*†** | -7.52 (6.48) | -0.07 (0.06) | -1.16 |
| **Covariates** |  |  |  |  |  |  |  |  |  |  |  |  |  |  |  |
| Age | -0.09 (0.10) | -0.06 (0.06) | -0.98 | 0.13 (0.09) | 0.09 (0.06) | 1.50 | -0.03 (0.08) | -0.02 (0.06) | -0.35 | -0.60 (0.12) | -0.29 (0.06) | **-5.11\*\*\*** | -0.31 (0.13) | -0.14 (0.06) | **-2.30\*†** |
| Sex | 2.18 (2.37) | 0.10 (0.11) | 0.92 | -1.81 (2.16) | -0.10 (0.12) | -0.84 | 5.64 (1.91) | 0.32 (0.11) | **2.95\*\*** | -1.86 (2.90) | -0.07 (0.11) | -0.64 | -0.96 (3.31) | -0.14 (0.12) | -0.29 |
| Education | 1.07 (0.35) | 0.17 (0.06) | **3.06\*\*** | 0.99 (0.32) | 0.19 (0.06) | **3.10\*\*** | 0.83 (0.28) | 0.16 (0.05) | **2.96\*\*** | 0.52 (0.43) | 0.07 (0.06) | 1.23 | -0.32 (0.49) | -0.04 (0.06) | -0.65 |
| NIHSS | -0.05 (0.20) | -0.02 (0.06) | -0.26 | -0.27 (0.18) | -0.09 (0.06) | -1.46 | -0.17 (0.16) | -0.06 (0.06) | -1.08 | -1.04 (0.25) | -0.24 (0.06) | **-4.21\*\*\*** | -0.45 (0.28) | -0.10 (0.06) | -1.61 |
| **Predictor** |  |  |  |  |  |  |  |  |  |  |  |  |  |  |  |
| 6-month cognitive impairment | -46.85 (6.75) | -0.40 (0.06) | **-6.94\*\*\*** | -13.56 (6.34) | -0.13 (0.06) | **-2.14\*†** | -24.33 (5.88) | -0.25 (0.06) | **-4.14\*\*\*** | -37.11 (8.25) | -0.26 (0.06) | **-4.50\*\*\*** | -30.35 (9.48) | -0.20 (0.06) | **-3.20\*\*** |
| **Covariates** |  |  |  |  |  |  |  |  |  |  |  |  |  |  |  |
| Age | 0.03 (0.10) | 0.02 (0.06) | 0.33 | 0.17 (0.09) | 0.12 (0.06) | 1.88 | 0.03 (0.09) | 0.02 (0.06) | 0.40 | -0.50 (0.12) | -0.24 (0.06) | **-4.24\*\*\*** | -0.22 (0.13) | -0.10 (0.06) | -1.64 |
| Sex | 1.32 (2.32) | 0.06 (0.11) | 0.57 | -2.17 (2.16) | -0.12 (0.12) | -1.00 | 4.80 (2.01) | 0.27 (0.11) | **2.39\*†** | -2.46 (2.82) | -0.09 (0.11) | -0.87 | -1.33 (3.25) | -0.05 (0.12) | -0.41 |
| Education | 0.85 (0.35) | 0.14 (0.06) | **2.44\*** | 0.94 (0.32) | 0.18 (0.06) | **2.90\*\*** | 0.84 (0.30) | 0.16 (0.06) | **2.79\*\*** | 0.23 (0.42) | 0.03 (0.06) | 0.55 | -0.58 (0.48) | -0.07 (0.06) | -1.20 |
| NIHSS | -0.18 (0.19) | -0.05 (0.06) | -0.94 | -0.33 (0.18) | -0.11 (0.06) | -1.85 | -0.40 (0.17) | -0.14 (0.06) | **-2.43\*†** | -1.01 (0.23) | -0.24 (0.05) | **-4.35\*\*\*** | -0.39 (0.27) | -0.09 (0.06) | -1.47 |

Hierarchical multivariable regressions were conducted for each SIS subscale using the proportion of tasks impaired on the OCS, both acutely and at 6 months post-stroke, while controlling for the same covariates. SIS: Stroke Impact Scale; OCS: Oxford Cognitive Screen; NIHSS: National Institute for Health Stroke Severity.
\*\*\* $p < 0.001$ , \*\* $p < 0.01$ , \* $p < 0.05$ , † No longer significant after correcting for multiple comparisons.

At 6 months, increased severity of cognitive impairment significantly predicted lower SIS composite scores after adjusting for demographics and stroke severity (*Adj R^2^*=0.15*, p*<0.001) (figure 2; table S2). Again, accounting for covariates, increased severity of 6-month cognitive impairment was also associated with poorer self-reported functioning in each of the SIS subscales: memory and thinking problems (Adj R^2^=0.19, *p*<0.001), mood and emotional difficulties (Adj R^2^=0.07, *p*=0.033), communication and language abilities (Adj R^2^=0.13, *p*<0.001), ADLs (Adj R^2^=0.21, *p*=0.001), and participation (Adj R^2^=0.05, *p*=0.002). Associations between severity of impairment and SIS subscale scores are shown in figures 2 and S1.

**Figure 2.**
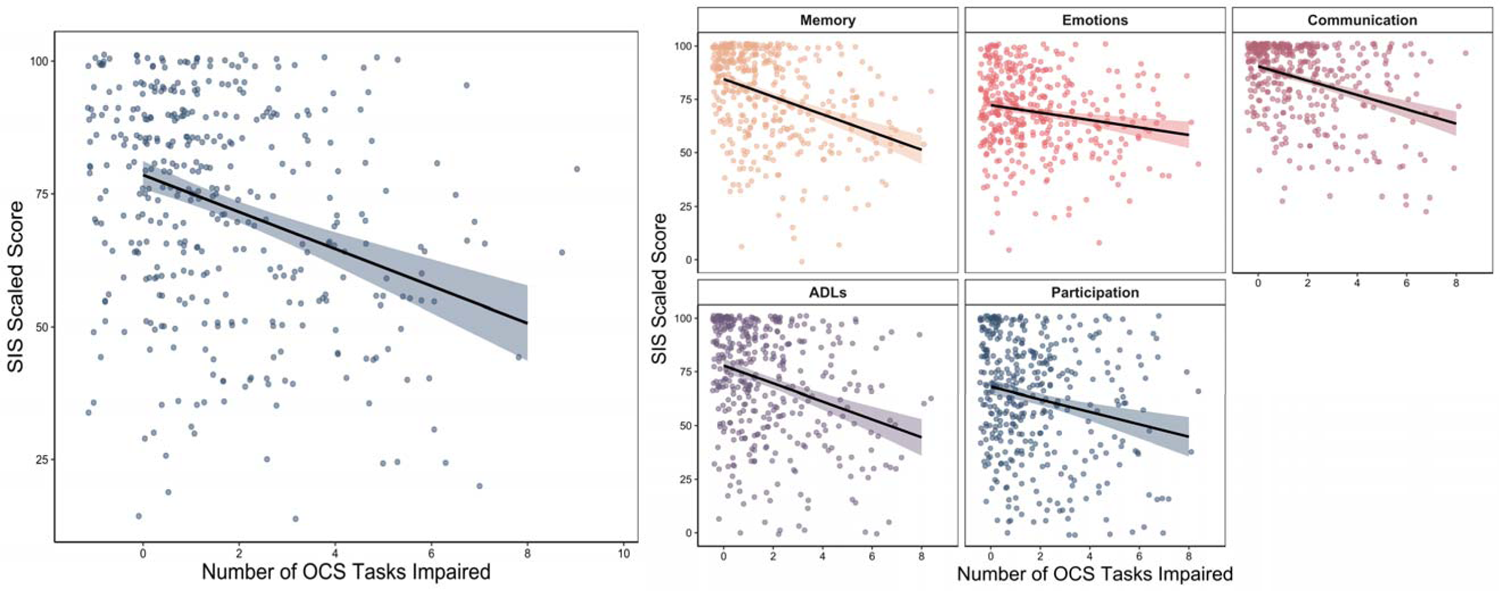
The association between severity of 6-month cognitive impairment and SIS scores.

### Acute and 6-month cognitive impairment and mood at 6 months

An overview of the regression results is shown in table S3. After adjusting for demographics and stroke severity, increased severity of acute cognitive impairment was associated with greater levels of self-reported depression (Adj *R*^2^=0.02, *p*=0.021), but not anxiety (Adj *R*^2^=0.03, *p*=0.174). Only lower age was a unique correlate of anxiety (Adj *R*^2^=0.03, *p*=0.027). Similarly, severity of cognitive impairment at 6 months was associated with greater levels of depression (Adj *R*^2^=0.05, *p*<0.001), but not anxiety (Adj *R*^2^=0.03, *p*=0.129) after adjustment. Again, only age was associated with greater levels of self-reported anxiety (Adj *R*^2^=0.03, *p*=0.015). The association between severity of impairment and HADS-Depression and HADS-Anxiety scores is visualised in figure S3.

### Acute and 6-month domain-specific cognitive impairments and composite SIS score

After adjusting for age, sex, education and stroke severity, acute impairments in memory (Adj R^2^=0.13, *p*<0.001), attention (Adj R^2^=0.10, *p*=0.002), and number processing (Adj R^2^= 0.08, *p*=0.016) were significantly associated with poorer overall self-reported function on the SIS composite score (though number processing did not survive correction for multiple comparisons). In contrast, impairments in language (Adj R^2^=0.07, *p*=0.05), executive function (Adj R^2^=0.06, *p*=0.624), and praxis (Adj R^2^=0.06, *p*=0.947) were not associated with SIS composite scores.

After controlling for covariates, 6-month impairments in language (Adj R^2^=0.13, *p*<0.001), memory (Adj R^2^=0.10, *p*=0.001), number processing (Adj R^2^=0.08, *p*=0.006) and praxis (Adj R^2^=0.06, *p*=0.045) domains were significantly associated with worse overall self-reported function on the SIS composite score (praxis no longer significant after multiple comparison correction). Executive dysfunction (Adj R^2^=0.05, *p*=0.459) and attention impairments (Adj R^2^=0.07, *p*=0.097) did not significantly contribute to SIS composite scores. An overview of associations between domain-specific cognitive function and 6-month SIS composite score is shown in table 3 and figure S1.

**Table 3.**
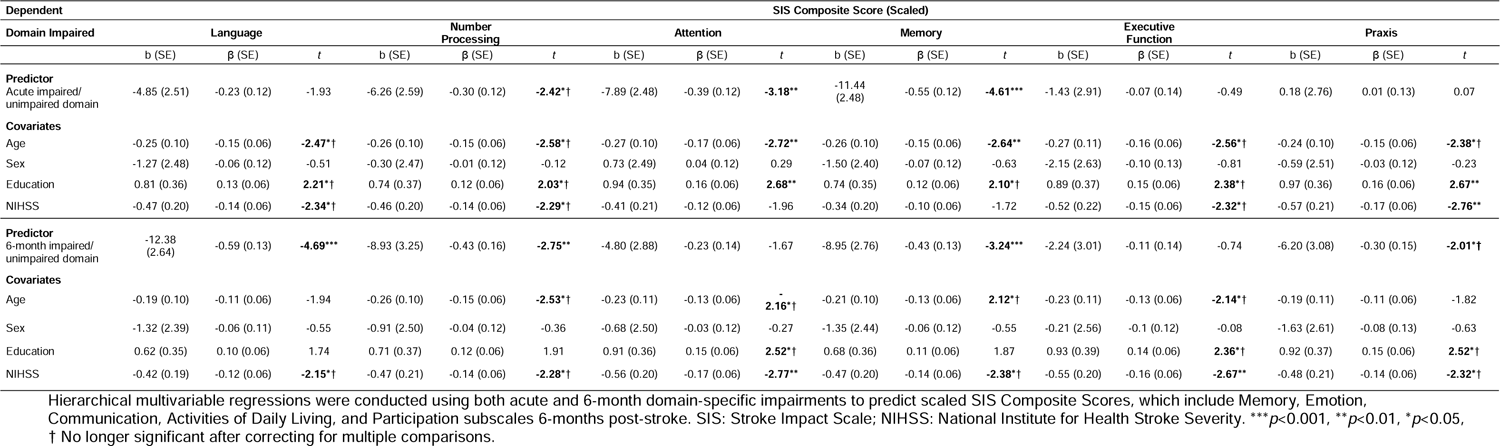
Associations between acute and 6-month domain-specific cognitive function and 6-month SIS composite score.

## DISCUSSION

This study sought to examine the relationship between acute and 6-month cognitive impairment (global severity and domain-specific), and overall quality of life 6 months post-stroke. Among 430 stroke survivors, highest self-reported functioning 6 months post-stroke was within the communication SIS subscale, followed by memory, ADLs, emotion, and participation. Severity of both acute and 6-month cognitive impairment was associated with lower 6-month SIS composite scores independent of age, sex, education, and stroke severity. When further explored, this association was present across all five individual SIS subscales, with the exception of acute cognitive impairment and the participation subscale. Severity of cognitive impairment at both timepoints was also associated with greater levels of self-reported depression, but not anxiety. Domain-specific cognitive impairments acutely in memory and attention and 6-month impairments in language, number processing and memory, were most strongly associated with worse overall 6-month SIS scores. Regarding prevalence of cognitive impairment, nearly all participants were impaired on at least one cognitive task on the OCS acutely. At 6 months follow-up, prevalence of domain-specific impairment had decreased across all domains, though two-thirds were impaired on at least one task.

At 6 months follow-up, participants reported poorest functioning in participation on average, followed by emotional functioning. This is in line with previous findings of SIS participation and emotional functioning being the most impacted subscale,^35,36^ and communication and memory perceived as least impacted subscales at both 3 months and one year.^36^ Emotional functioning and reduced participation are likely to co-occur following stroke with possible reciprocal effects.^37^ However, Lee and colleagues^37^ found general emotion measured by the SIS was not a significant contributor to post-stroke participation in the short or long-term, and instead found specific emotional symptoms (depression, anxiety, and apathy) to be strongly associated with post-stroke participation. This suggests treatments targeting specific symptoms rather than emotional status more generally may be useful in participation interventions. Additionally, as emotional functioning and participation are among the top reported long-term unmet care needs of stroke patients,^7,9^ our findings support prioritizing these areas of need within the care pathway.

Previous research has identified that global cognitive impairment after stroke is associated with depression at 6 months.^4,38–40^ Using the stroke-specific OCS, our study replicated these findings to show that increased severity of cognitive impairment, both acutely and at 6-months follow-up, was associated with heightened depressive symptomatology at 6 months. The variance explained in both acute and 6-month models was modest (5%), aligning with previous studies.^41^ Using unstandardized coefficients (b values), we found a small change in depressive symptoms acutely (b=0.22) and a moderate change at 6 months (b=0.50) for a one-unit increase in cognitive impairment.^42^ These findings underscore the relevance of considering cognitive impairment in understanding post-stroke depressive symptomatology. Notably, as a Minimal Clinically Important Difference (MCID) for the HADS in the stroke population remains unestablished, future research in this area is warranted. Determining an MCID specific to stroke patients could enhance our understanding of the clinical significance of cognitive-depressive associations and inform targeted interventions for this population. Our findings also suggest an additive effect of cognitive deficits on the risk of post-stroke depression, or alternatively, altered mood post-stroke may hinder cognitive recovery or impact performance on cognitive tests.

Interventions targeting cognitive recovery or depressive mood may positively impact both cognition and mood. In contrast, a greater severity of cognitive impairment was not associated with anxiety as seen in some,^4^ but not all previous studies.^24^ Younger age was the only factor linked to anxiety in our study, aligning with previous reports of three to four-fold increase in anxiety symptoms in those younger than 65 years.^43,44^ Younger individuals, particularly of working age, may experience more anxiety symptoms due to stroke threatening survival, independence, loss of identity, and ability to participate in occupational and social activities.^45^ Collectively, our findings suggest variations in the depression-cognition relationship among stroke survivors, indicating potential specific impairments or combinations that elevate depression risk. Saxena et al.^46^ suggest that improving depressive symptoms in stroke patients may enhance functional recovery, though limited by cognitive impairment. Despite a relatively weak relationship between cognition and depression observed, treatment implications exist. The reported low value might be influenced by common method variance, where objective measures like the Oxford Cognitive Screen lower correlations with self-report scales such as the HADS. For the anxiety-cognition relationship, given other research has not found cognition to be a predictor (e.g., Chun et al.^45^), it is less likely that we have underestimated the relationship.

Results underscore the need for future studies to identify potential vulnerable subpopulations for targeted early interventions, emphasizing the importance of individualized care, even with relatively low correlation coefficients. Clinical significance is highlighted by Obaid et al.^39^ linking early post-stroke cognitive impairment to increased risk of mortality, physical dependency, depression, and institutionalization at one and five years.

This study demonstrated a significant association between the severity of both acute and 6-month cognitive impairment and poorer functioning in SIS composite scores and subscales. These associations remained significant after controlling for demographic factors and stroke severity, contributing to the literature where such comparisons were scarcely reported.^6^ Our findings indicate that acute cognitive impairment explained approximately 10% of the variance in 6-month SIS composite scores, with adjusted R-squared values ranging from 7% in emotions to 21% in communication. Similarly, increased severity of 6-month cognitive impairment explained 15% of the variance in 6-month SIS scores, with adjusted R-squared values ranging from 5% in participation to 21% in activities of daily living. These effect sizes align with Verhoeven et al.^41^ who reported a range of percentages of variance explained by total cognition scores at one-year post-stroke, such as 8% for life satisfaction, 19% for functional independence, and 40% for participation. Notably, the observed differences in adjusted R-squared values across SIS subdomains may reflect the varying cognitive demands associated with different functional domains. For instance, participation, requiring more complex cognitive abilities and potentially being vulnerable to non-cognitive determinants such as mood,^37^ demonstrated lower adjusted

R-squared values compared to activities of daily living. Existing literature often links cognition more strongly to basic ADLs than to complex ADLs,^6^ which could contribute to the observed variations. Despite the modest variance explained by conventional standards, the consistent associations across diverse SIS subdomains underscore the impact of cognitive impairment on various aspects of post-stroke functioning. Additionally, our findings align with Guidetti et al.^35^ who suggested changes of approximately 10-15 points in SIS domain scores may be considered clinically meaningful. Notably, the increased variance explained in 6-month SIS scores suggests a potentially more influential role of the severity of cognitive impairment at 6 months in determining post-stroke quality of life compared to acute impairment. Our findings also indicate there may be a certain threshold of cognitive impairment that must be surpassed in order to affect function and routine daily activities frequently or evident enough to be acknowledged and reported by the patient. Patients are typically discharged after short acute care and often expected to return to their previous lives without major difficulties, particularly in the absence of physical disability.^47^ Further emphasis should be placed on the importance of follow-up cognitive screening post-stroke. This will identify individuals with more subtle limitations in need of individualized support after stroke to improve quality of life.

Few studies have considered all domains comprehensively when assessing the impact of cognitive function on quality of life. However, a variety of individual cognitive domain impairments have been reported to contribute to poor functional outcome and quality of life at or beyond one year, such as memory,^1,12,48^ attention,^1,49^ executive functions,^1,12,48^ language,^12,48^ and visuospatial ability.^1,12,49^ In our study, domain-specific deficits in attention (adjR=0.10) and memory (adjR=0.13) acutely, and number processing, memory and language at 6 months (adjR=0.08-0.13), were shown to be associated with poorer perceived quality of life 6 months post-stroke. This emphasizes the enduring impact of acute cognitive challenges and suggests that the recovery, or lack thereof, in these particular domains during the subacute phase directly shapes individuals’ quality of life at that stage.

Acute attention deficits, although often resolving within the first few months post-stroke, significantly influence functional outcomes at 6-months.^50^ Number processing deficits also tend to recover substantially by 6-months.^51,52^ However, individuals with subacute deficits in number processing, which contains OCS tasks which are heavily loaded on working memory (calculations and arithmetic),^53^ often exhibit multi-domain impairments, increasing the risk of poorer overall quality of life. Language domain deficits, with comparatively low recovery rates,^11^ impact instrumental ADL and social participation, (e.g., reduced confidence in speaking abilities and difficulty returning to employment) compared with age-matched controls.^54^ However, conflicting evidence surrounds whether adults with post-stroke aphasia experience worse quality of life compared to non-aphasic stroke counterparts.^54–56^ Evidence suggests follow-up time and adaptation strategies are important factors in determining functional outcome as those with aphasia for more than 6 months tend to report improved quality of life than recently aphasic individuals.^55^ Interestingly, the temporal relationship between specific deficits and quality of life does not seem to apply to memory impairment post-stroke, suggesting the critical role of memory in determining quality of life prognosis. Persistent and acquired memory deficits may be linked to pre-stroke neurodegeneration, with lasting implications on daily functioning, mood, and compensating strategies.^57^ Collectively, findings highlight the dynamic interplay between a variety of domain-specific cognitive impairments and post-stroke quality of life. Recognizing these relationships and enabling early detection of impairments, particularly memory deficits, could inform tailored rehabilitation strategies, optimizing outcomes and lessening the long-term impact on stroke survivors’ quality of life.

A notable finding within the present study is executive dysfunction was not associated with overall poorer self-reported quality of life. This differs from recent reports where executive dysfunction was strongly related to post-stroke functional outcomes, including the Modified Rankin Scale and IADLs, at three months.^58^ These disparities may be explained by differences in time point of assessment after stroke, though could also be due to differences in how executive function was characterized and assessed. It may also represent a de-coupling of executive function abilities as stroke survivors adapt to the long-term psychological impact of stroke. For example, cognitive impairment (e.g., executive dysfunction) has been shown to be associated with reduced participation,^13^ though this relationship was found to be moderated by length of follow-up period, with shorter follow-up time demonstrating a stronger association than in longer follow-up.^6^ Additionally, executive dysfunction alone may manifest in more nuanced changes to an individual’s daily function compared to a language or memory impairment for example, and therefore the individual may not perceive to be as severely impacted by stroke.^59^ This notion is supported by the association found between language, memory, and worse quality of life, perhaps indicating impairments more outwardly apparent to the patient may result in worse self-perceived function. Further examination using in-depth neuropsychological characterization of executive function at various timepoints post-stroke and how it relates to functional outcome and overall quality of life is warranted.

The key strengths of this study are the consecutive recruitment of an inclusive sample with a balance of both minor and major stroke, as well as the use of the stroke-specific OCS to assess impairment in individual cognitive domains both acutely and at 6 months. However, the results of this study should be considered in light of several limitations. There was considerable attrition from acute to 6-month follow-up, which could introduce a degree of attrition bias with more severely impaired individuals being lost to follow-up. Despite best efforts to retain participants, attrition in representative clinical samples is often unavoidable due to high rates of death and severe illness, although many participants also declined follow-up or were unable to be contacted. Secondly, the nature of cognitive studies requires a sufficient level of comprehension in order to consent, which may preclude individuals with most severe stroke from participating. Third, though the SIS is stroke-specific, it is a structured self-report questionnaire and cognitive impairment may interfere with the ability to complete a questionnaire accurately. Nevertheless, patient-report questionnaires are the optimal way to evaluate the individual’s perspective not obtained through clinician or informant report. Lastly, pre-stroke cognitive status, depression, and anxiety data were not available for the current sample.

## Conclusion

This study uniquely employs a domain-specific cognitive screening tool to investigate the association between cognition and multiple functional outcomes. At 6 months, stroke survivors reported decreased levels of activities of daily living, participation, and emotional functioning, communication and memory, and cognitive impairments were prevalent across all six OCS domains. The severity of cognitive impairment, particularly subacutely, was found to be significantly associated with all five SIS components of functional outcome, as well as depressive symptomatology. Memory deficits, both acutely and at 6-months, appear to be a key factor influencing poorer quality of life at 6 months. Collectively, these findings highlight the need to identify domain-specific impairments early to differentiate individuals who are at increased risk of functional limitations, especially reduced emotional functioning and levels of participation.

## Data Availability

The participants of this study did not give written consent for their individual data to be shared publicly, so due to the sensitive nature of the research supporting data is not available.

## Acknowledgements

This study was funded by the Stroke Association (SA PPA 18/100032). ND is supported by an NIHR Advanced Fellowship (NIHR302224). STP is supported by the NIHR Oxford BRC. This study was supported by the NIHR Clinical Research Network. The views expressed are those of the authors and not necessarily those of the NHS, NIHR or Department of Health. We thank all participants and members of the Oxford Translational Neuropsychology Group for contributions to recruitment/testing.

## Declaration of Interest

ND is a developer of the Oxford Cognitive Screen but does not receive renumeration for its use. STP has received honoraria from Trondheim, Sydney and LaTrobe universities and royalties from Oxford University Press and Cambridge University Press.

## Notes

### Author Declarations

National Research Ethics Committee (UK) gave approval for this work (references 14/LO/0648 and 18/SC/0550)

