## Supplemental Material for "Domain-specific Cognitive Impairments, Mood and Quality of Life 6 Months After Stroke"

### Supplementary Materials

#### Supplemental methods – Patient cohort recruitment, attrition, and follow-up.

**Figure S1.** Flow chart of patient cohort from baseline to 6-month follow-up.

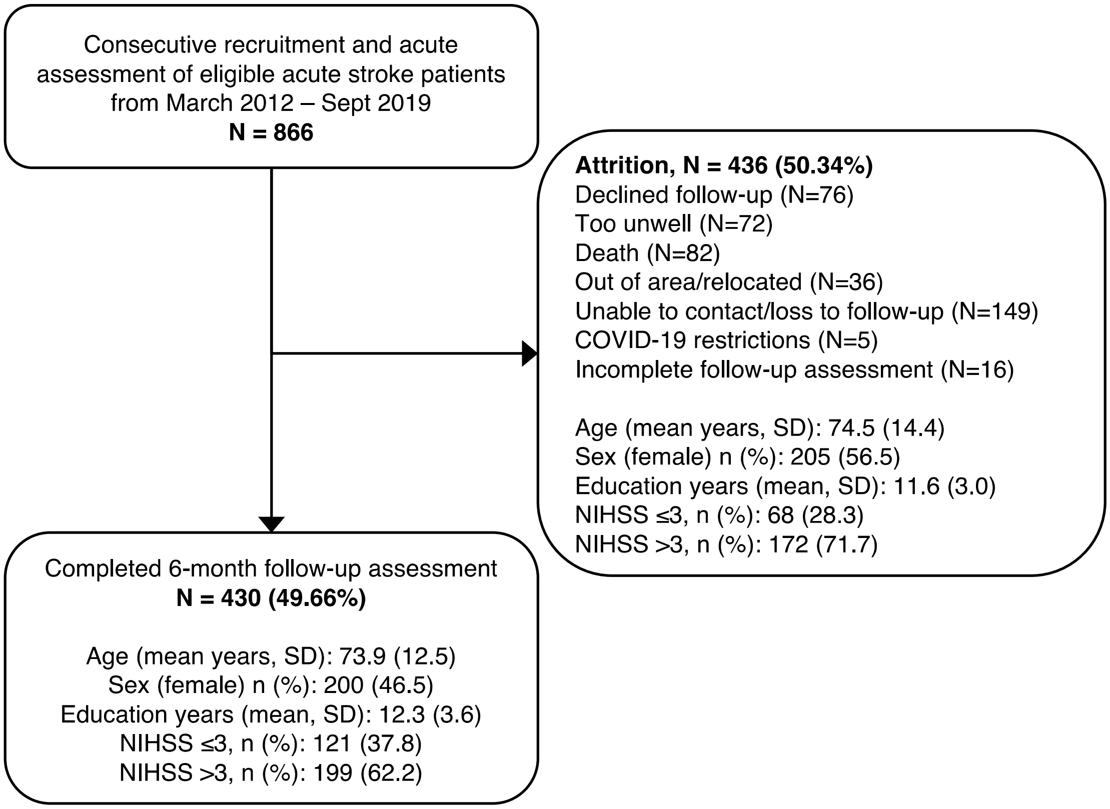

**Supplemental methods –** Oxford Cognitive Screen (OCS)

The OCS is a cognitive screening tool designed specifically for the stroke population, assessing common impairments whilst reducing potential confounds and being inclusive for patients with aphasia and neglect.[31] A description of the specific tasks and corresponding normative data and validation is detailed elsewhere.[31] The OCS takes approximately 20 minutes to administer. Cognitive data was included in analysis if participants were able to complete at least 70% of OCS tasks, allowing for sufficient evaluation of each cognitive domain. Participants who attempted but were unable to complete a task due to an impairment (as judged by the assessor) were given the minimum score and considered impaired on that task (e.g., in cases of aphasia, picture naming was often scored zero, while those with hemiplegia were able to use their non-dominant arm for the praxis task). In cases where participants were unable to complete a task due to time, other administrative constraints, or severe blindness, data from tasks that were not administered were not considered when classifying domain impairments.

**Supplemental results – 1:** Group differences between those who completed 6-month follow up (N = 430) and those who did not (N = 436).

A total of 866 stroke patients were recruited and assessed acutely, 430 (49.7%) of which completed a follow-up assessment at 6 months (Figure 1). Age ($t$ = 0.677, p = 0.498), sex ($X^2$ = 0.007, p = 0.935), education ($t$ = 2.480, p = 0.132), and handedness ($X^2$ = 0.00, p = 0.993) did not statistically differ between those who were re-assessed and those who were not. Stroke severity (NIHSS) did differ between groups ($X^2$ = 5.096, p = 0.002), with a greater proportion of more severe strokes (NIHSS >3) occurring in the group that was not re-assessed (71.7%) compared to those who were (62.2%). Additionally, patients who were not re-examined demonstrated a significantly ($p$ <0.05) greater proportion of impairment across all domains except attention, including language (58.3% vs 45.2%, p <0.001), executive function (36.4% vs 29.3%, p=0.046), memory (56.0% vs 39.9%, p <0.001), number (49.0% vs 41.7%, p=0.007) and praxis (39.1% vs 26.7%, p <0.001). However, overall, there was also a greater proportion of patients who had no impairments compared to those who completed a 6-month assessment (14.5% vs 1.6%). For the purposes of this paper, the main focus was on how longer-term impairments impact quality of life; thus, only patients who completed follow-up were included in analyses.

**Table S1:** Descriptive statistics for the Oxford Cognitive Screen, the Stroke Impact Scale and the Hospital Anxiety and Depression Scale (*N* = 430).

| | **Acute** | **6 months** |
|---|---|---|
| *OCS domain impairments, n (%)* | | |
| **Language** | 195/429 (45.5) | 138/430 (32.1) |
| Picture naming | 139/429 (32.4) | 78/428 (18.2) |
| Semantic understanding | 42/427 (9.8) | 11/252 (4.4) |
| Sentence reading | 136/419 (32.5) | 88/422 (20.9) |
| **Spatial attention** | 177/391 (45.3) | 101/416 (24.3) |
| Egocentric attention | 118/430 (27.4) | 54/414 (13.0) |
| Allocentric attention | 107/391 (27.4) | 101/416 (24.3) |
| **Executive function** | 113/380 (29.7) | 111/413 (26.9) |
| **Memory** | 170/428 (39.7) | 137/430 (31.9) |
| Orientation | 90/392 (23.0) | 72/426 (16.9) |
| Verbal memory | 106/431 (24.6) | 78/417 (18.7) |
| Episodic memory | 77/389 (19.8) | 54/416 (13.0) |
| **Number processing** | 178/428 (41.6) | 85/420 (20.2) |
| Calculations | 72/426 (16.9) | 39/417 (9.4) |
| Writing | 161/393 (41.0) | 66/409 (16.1) |
| **Praxis** | 112/419 (25.9) | 79/403 (19.6) |
| *SIS subscale scores, mean (SD), [Min-Max]* | | |
| **Memory** | | 77.1 (21.4) [0 – 100] |
| **Emotions** | | 69.2 (18.4) [5.6 – 100] |
| **Communication** | | 84.4 (18.2) [14.3 – 100] |
| **Activities of daily living** | | 70.4 (26.3) [0 – 100] |
| **Participation** | | 62.8 (27.3) [0 – 100] |
| *HADS subscale scores (mean, SD) and cases, n (%)* | | |
| **Depression** subscale score | | 5.4 (4.1) |
| Possible depression cases (score >7) | | 101/430 (23.5) |
| Probable depression cases (score >10) | | 61/430 (14.2) |
| **Anxiety** subscale score | | 5.7 (4.3) |
| Possible anxiety cases | | 124/430 (28.8) |
| Probable anxiety cases | | 78/430 (18.1) |

Oxford Cognitive Screen (OCS) domain impairment defined by an impairment on any within-domain subtest; Stroke Impact Scale (SIS) scaled scores range between 0 – 100 with higher scores indicating better self-reported function; Hospital Anxiety and Depression Scale (HADS) observed scores range from 0 – 19 for depression and 0 – 20 for anxiety in this cohort, higher scores indicate increased depression/anxiety symptomatology, possible depression/anxiety: scores >7, Probable depression/anxiety: scores >10.

**Table S2.** Association between severity of acute and 6-month cognitive impairment and SIS composite score.

| Dependent Variable | SIS Composite Score (Scaled) | | |
|---|---|---|---|
| **Predictors** | *b (SE)* | β (SE) | *t* |
| **Acute cognitive impairment** | -16.30 (4.76) | -0.21 (0.06) | **-3.42**** |
| *Covariates* | | | |
| Age | -0.25 (0.10) | -0.15 (0.06) | **-2.45*** |
| Sex | -0.54 (2.44) | -0.03 (0.12) | -0.22 |
| Education | 0.76 (0.36) | 0.13 (0.06) | **2.12*** |
| NIHSS | -0.31 (0.21) | -0.09 (0.06) | -1.49 |
| **6-month cognitive impairment** | -37.74 (6.88) | -0.33 (0.06) | **-5.48***** |
| *Covariates* | | | |
| Age | -0.13 (0.10) | -0.08 (0.06) | -1.33 |
| Sex | -1.30 (2.36) | -0.06 (0.11) | -0.55 |
| Education | 0.51 (0.35) | 0.08 (0.06) | 1.46 |
| NIHSS | -0.33 (0.19) | -0.10 (0.06) | -1.69 |

Full hierarchical multivariable regressions with *b*-values, beta values (β), standard errors (*SE*) and *t*-values (*t*). SIS: Stroke Impact Scale; OCS: Oxford Cognitive Screen; NIHSS = National Institute for Health Stroke Severity.
***$p$ <0.001, **$p$ <0.01, *$p$ <0.05

**Figure S1.** Point-biserial correlations with alpha-adjusted significance levels between Oxford Cognitive Screen (OCS) domains and Stroke Impact Scale (SIS) subscale.

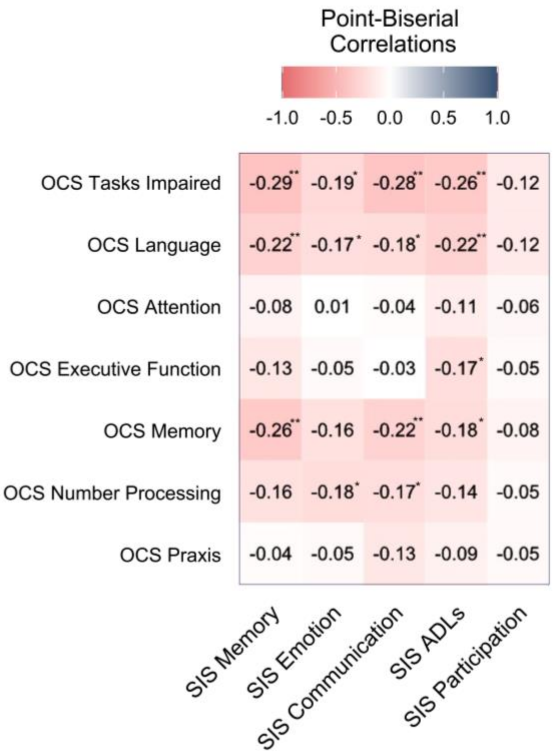


Note for each SIS subscale correlation, the item with the strongest loading to each subscale total (as in Jenkinson and colleagues)[33] was used so as to be comparable to the main text regression analyses. **$p$ < 0.01, * $p$ < 0.05.

**Table S3.** Association between severity of cognitive impairment and emotional status.

| Dependent Variable | HADS Depression | | | HADS Anxiety | | |
|---|---|---|---|---|---|---|
| **Predictors** | *b* (SE) | β (SE) | *t* | *b* (SE) | β (SE) | *t* |
| **Acute cognitive impairment** | 0.22 (0.09) | 0.15 (0.06) | **2.33*†** | 0.14 (0.10) | 0.09 (0.06) | 1.36 |
| *Covariates* | | | | | | |
| Age | 0.02 (0.02) | 0.06 (0.06) | 1.04 | -0.05 (0.02) | -0.14 (0.06) | **-2.22*** |
| Sex | -0.20 (0.49) | -0.05 (0.12) | -0.41 | 0.46 (0.53) | 0.11 (0.12) | 0.87 |
| Education | -0.07 (0.07) | -0.06 (0.06) | -0.91 | -0.14 (0.08) | -0.11 (0.06) | -1.86 |
| NIHSS | 0.03 (0.04) | 0.04 (0.06) | 0.66 | 0.02 (0.04) | 0.03 (0.06) | 0.42 |
| **6-month cognitive impairment** | 0.50 (0.13) | 0.24 (0.06) | **3.93*** ** | 0.21 (0.14) | 0.10 (0.06) | 1.52 |
| *Covariates* | | | | | | |
| Age | 0.01 (0.02) | 0.02 (0.06) | 0.30 | -0.05 (0.02) | -0.15 (0.06) | **-2.44*** |
| Sex | -0.09 (0.48) | -0.02 (0.12) | -0.18 | 0.51 (0.53) | 0.12 (0.12) | 0.98 |
| Education | -0.03 (0.07) | -0.02 (0.06) | -0.38 | -0.13 (0.08) | -0.11 (0.06) | -1.68 |
| NIHSS | 0.03 (0.04) | 0.05 (0.06) | 0.76 | 0.03 (0.04) | 0.04 (0.06) | 0.60 |

Full hierarchical multivariate regressions with *b*-values (*b*), Beta values (β) standard errors (*SE*) and *t*-values (*t*). Two regressions using proportion of tasks impaired on the OCS acutely and at 6 months post-stroke with the same covariates were conducted with each HADS subscale listed above. HADS: Hospital Anxiety and Depression Scale; NIHSS: National Institute for Health Stroke Severity.
*** $p<0.001$, ** $p<0.01$, *$p<0.05$, † No longer significant after correcting for multiple comparisons.

**Figure S3**. Association between severity of cognitive impairment and emotional status.

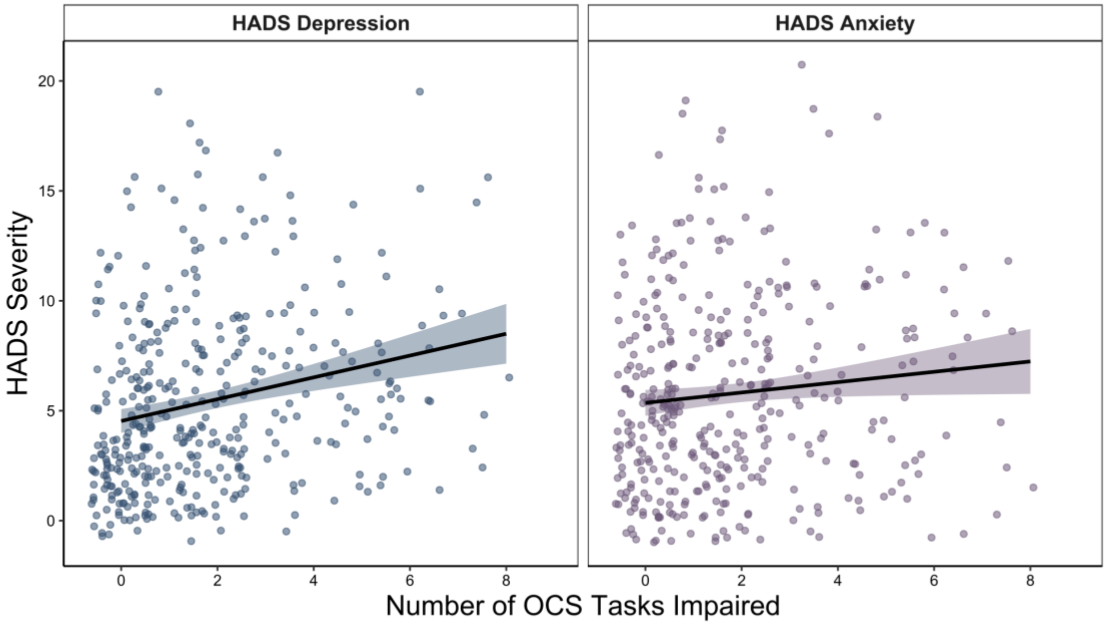


The association between severity of 6-month cognitive impairment and depression ($r$ =0.05, $p$<0.01) **(left)**, and anxiety ($r$=0.03, $p$<0.126) **(right)**. Severity of impairment defined by proportion of OCS tasks impaired. HADS: Hospital Anxiety and Depression Scale; OCS: Oxford Cognitive Screen. Note jitter has been applied to data points for visualization purposes only.
